# Comorbidities and Disparities in Outcomes of COVID-19 Among Black and White Patients

**DOI:** 10.1101/2020.05.10.20090167

**Authors:** Ahmad Khan, Arka Chatterjee, Shailendra Singh

## Abstract

Initial surveillance data suggests a disproportionately high number of deaths among Black patients with COVID-19. However, high-risk comorbidities are often over-represented in the Black population, and understanding whether the disparity is entirely secondary to them is essential. We performed a retrospective cohort study using real-time analysis of electronic medical records (EMR) of patients from multiple healthcare organizations in the United States. Our results showed that Black patients with COVID-19 have a significantly higher risk of mortality, hospitalization, and invasive mechanical ventilation compared to White patients. The incremental risk of poor outcomes in Blacks persists despite accounting for a higher prevalence of comorbidities. This may point to the disparities in socioeconomic determinants of health affecting Blacks and the need for an improvement in the care of this vulnerable population.

## Main

### Background

Coronavirus disease 2019 (COVID-19) caused by severe acute respiratory syndrome coronavirus 2 (SARS CoV-2) infection has emerged as unprecedented global health, economic, and social crisis. More than 2 million COVID-19 cases and 160,000 deaths have been reported worldwide as of April 19, 2020 [1]. Initial surveillance data from 14 states suggests a disproportionately high number of cases and deaths for Black or African Americans [2], However, it has also been observed that comorbid conditions such as hypertension (HTN), diabetes mellitus (DM), and cardiovascular diseases are associated with worse outcomes with COVID-19 infection[3, 4], These comorbidities are often over-represented in Black communities[5]. Thus, an understanding of the outcomes among Black patients with COVID-19 in the milieu of these comorbidities is required. We attempt to provide insight into this with a propensity-matched analysis of a large cohort of Black and white COVID 19 patients.

## Methods

### Study Design and Data Source

This was a retrospective cohort study conducted using TriNetX (Cambridge, MA, USA), a global federated health research network providing real-time access to electronic health records (EHR) of more than 40 million patients from 34 member healthcare organizations (HCOs). TriNetX provides access to HIPPA compliant de-identified longitudinal clinical data to member Health Care Organizations (HCOs) in a real-time fashion on its cloud-based platform. The de-identified clinical data such as diagnoses, procedures, medications, laboratory values, and genomic information are aggregated directly from the EHR systems of the member HCOs continuously. Both the patients and HCO’s as data sources stay anonymous. Member HCO’s include a mix of inpatient, outpatient, and specialty care services and provide care to a diverse patient population from different age groups, ethnicity, income levels, and geographical region. TriNETX recently fast-tracked data inflow to incorporate COVID-19 specific diagnosis and terminology following the World Health Organization (WHO) and Centers for Disease Control (CDC) COVID-19 criteria for coronavirus research. TriNetX data has also been granted a waiver from the Western IRB because it is a federated network, and only aggregated counts and statistical summaries of the de-identified information without any protected health information is received from participating HCO’s. Further details of the data source, data quality checks, mapping, and coding systems to present data are available in the Supplementary.

### Patient Selection

#### Selection of COVID-19 patients

We performed a real-time search query (April 21, 2020) on the research network to identify patients > 10 years with a diagnosis of COVID-19 between January 20, 2020, and April 21, 2020. The search criteria to identify potential COVID-19 patients was based on specific diagnosis and terminology recommended by the WHO and CDC. These codes included International Classification of Diseases, Ninth Revision and tenth Revision, Clinical Modification (ICD-10-CM) codes U07.1 (COVID-19, virus identified), B34.2 (Coronavirus infection, unspecified), B97.29 (Other coronavirus as the cause of diseases classified elsewhere) and J12.81 (Pneumonia due to SARS-associated coronavirus). Patients identified with diagnosis code 079.89 (Other specified viral infection) and Z03.818 (Encounter for observation for suspected exposure to other biological agents ruled out) were excluded. Only patients diagnosed with the codes, as mentioned earlier after January 20, 2020 (first confirmed case in the USA) were included. The B97.29 code was specifically included based on the recommendation from the general guidance of the ICD-10-CM Official Coding Guidelines released by the CDC on February 20, 2020. Similarly, U07.1 is the new specific code for a confirmed diagnosis of the COVID-19 with a positive COVID-19 test result starting April 1, 2020, as per the new CDC guidelines. The codes B34.2 and J12.81 were used more often before the CDC guidelines. Patients with ICD-9 code 079.89 (mapped to ICD-10 code B34.2 and B97.2) were excluded to reduce any false positive COVID-19 patients because this ICD-9 code can still be used occasionally as “catch-all’ code for more than 50 viral infections.

### Selection of Black and White Patient Cohort

Patient demographics, including race, and ethnicity, are coded to HL7 version 3 administrative standards. Races and Ethnicity are self-reported and categorized as per the United States (U.S) Census Bureau [6]. Federal standards acknowledge that race is a social-political construct, and self-identification with one more race categories is preferred to observer identification. The most recent United States Census officially recognized five racial categories (White, Black or African American, American and Alaska Native, Asian, and Native Hawaiian and Other Pacific Islander). We only included COVID-19 patients with races identified as Black or African American (Black cohort) and White American (White cohort) (Figure 1). We excluded patients with race identified as Asian, American Indian or Alaska Native, Native Hawaiian or other Pacific Islander, or with unknown race. The reporting of Ethnicity is separate from the race and can be categorized as Non-Hispanic or Latino and Hispanic or Latino as per the U.S census. Hispanics could report as any race [6]. We excluded patients identified as Hispanics from the white cohort for a more generalizable comparison between white and black patients.

**Figure 1:**
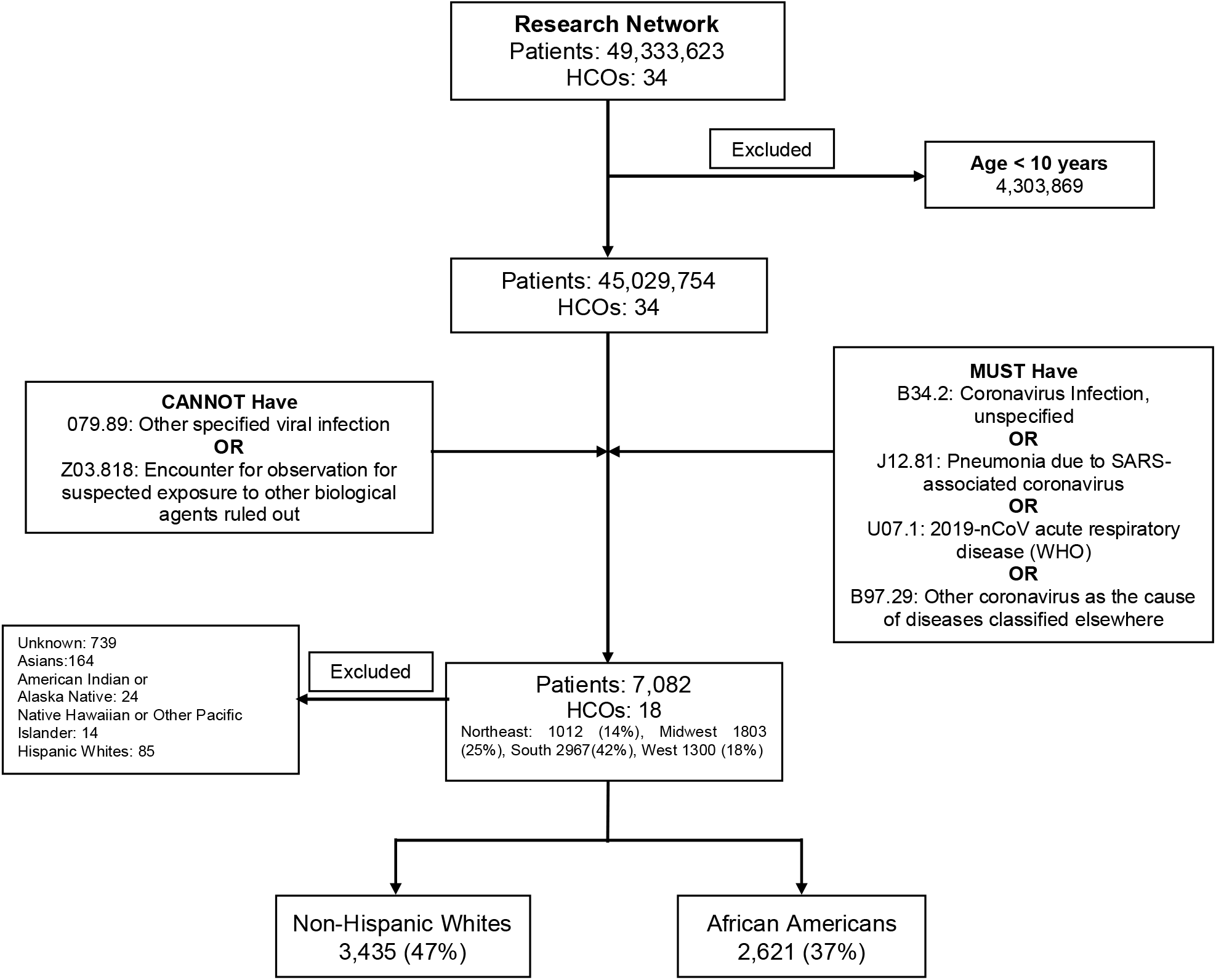
Flow chart showing the selection of African American and White COVID-19 patient cohorts.

### Definition of Variables and Outcomes

TriNetX has the capability of analyzing data based on a temporal relationship to the index event. The index event in our study was defined as the diagnosis of COVID-19. Baseline characteristics were estimated from any time before the index event. Presenting signs and symptoms were recorded from the time of index event up to two weeks before the index event. The outcomes were assessed from the index event up to 30 days after the index event. The primary outcome was the mortality in Black and white COVID-19 patients. Other outcomes of interest were mechanical ventilation, hospitalization, critical care services, development of acute respiratory distress syndrome (ARDS), septic shock, acute kidney injury (AKI), and continuous renal replacement therapy (CRRT) requirement. We also performed a subgroup analysis of mortality stratified by age into three sub-groups ≤49, 50-64, and ≥65 years. Subgroups were divided based on the mean age of the cohort and the risk of poor outcomes while obtaining a comparable sample size in each group. We also compared the laboratory findings after the diagnosis of COVID-19. Codes used to define outcomes (Supplementary Table 1), and laboratory values (Supplementary Table 2) can be found in the Supplementary.

### Statistical Analysis

We conducted all the statistical analyses using TriNetX, which has inbuilt analytics features. Descriptive statistics were reported as means, standard deviations for continuous variables, and total counts with frequencies for categorical variables. However, patient counts were obfuscated by rounding up to the nearest 10 in the analysis to safeguard the protected health information. We performed the inferential statistics by calculating the risk difference and risk ratio for the comparison of each outcome in both the groups. However, to adjust for the confounding factors, we performed 1:1 propensity score matching using a greedy nearest-neighbor matching algorithm with a caliper of 0.1 standard deviations. The outcomes are reported with and without propensity matching. An alpha of less than 0.05 was considered as the cutoff for statistical significance.

## Results

A total of 7,082 COVID-19 patients from 18 HCOs were identified. The mean age for these COVID-19 patients was 51.3 ±18.2 years, and the majority of them were females (60%). Out of these, 3,435 white patients and 2,621 black patients were included in the study (Figure 1).

Table 1 compares the characteristics between the Black and white COVID-19 patient cohorts. Patients in the Black cohort were older than as compared to the white cohort (mean age 54.1±16.8 vs. 50.9 ± 18.7, P<0.001). Black patients had higher BMI and high rates of hypertension, diabetes, chronic kidney disease, and heart failure. The prevalence of chronic respiratory disease (Asthma and COPD combined) was higher in white patients, while the rate of Ischemic heart disease was similar. Propensity matching of cohorts was performed accounting for age, gender, BMI, and major comorbidities described in Table 1. After propensity score matching, there were 1862 patients in each group, and the cohorts were well-balanced (Figure 2 and Table 1). Presenting clinical signs and symptoms are also described in Table 1. Laboratory findings in each group after the diagnosis of COVID-19 are described in Supplementary Table 3. Significantly higher creatinine (mg/dl) (2.13 ±2.51 Vs. 1.25 ±1.27, P <0.001), erythrocyte sedimentation rate (mm/hr) (81.04 ±34.95 Vs. 53.96 ±29.82, P <0.001), and C-Reactive Protein (mg/L) (101.31 ±107.22 Vs. 85.27 ±90.16, P: 0.004) levels were seen in black patients.

**Table 1:**
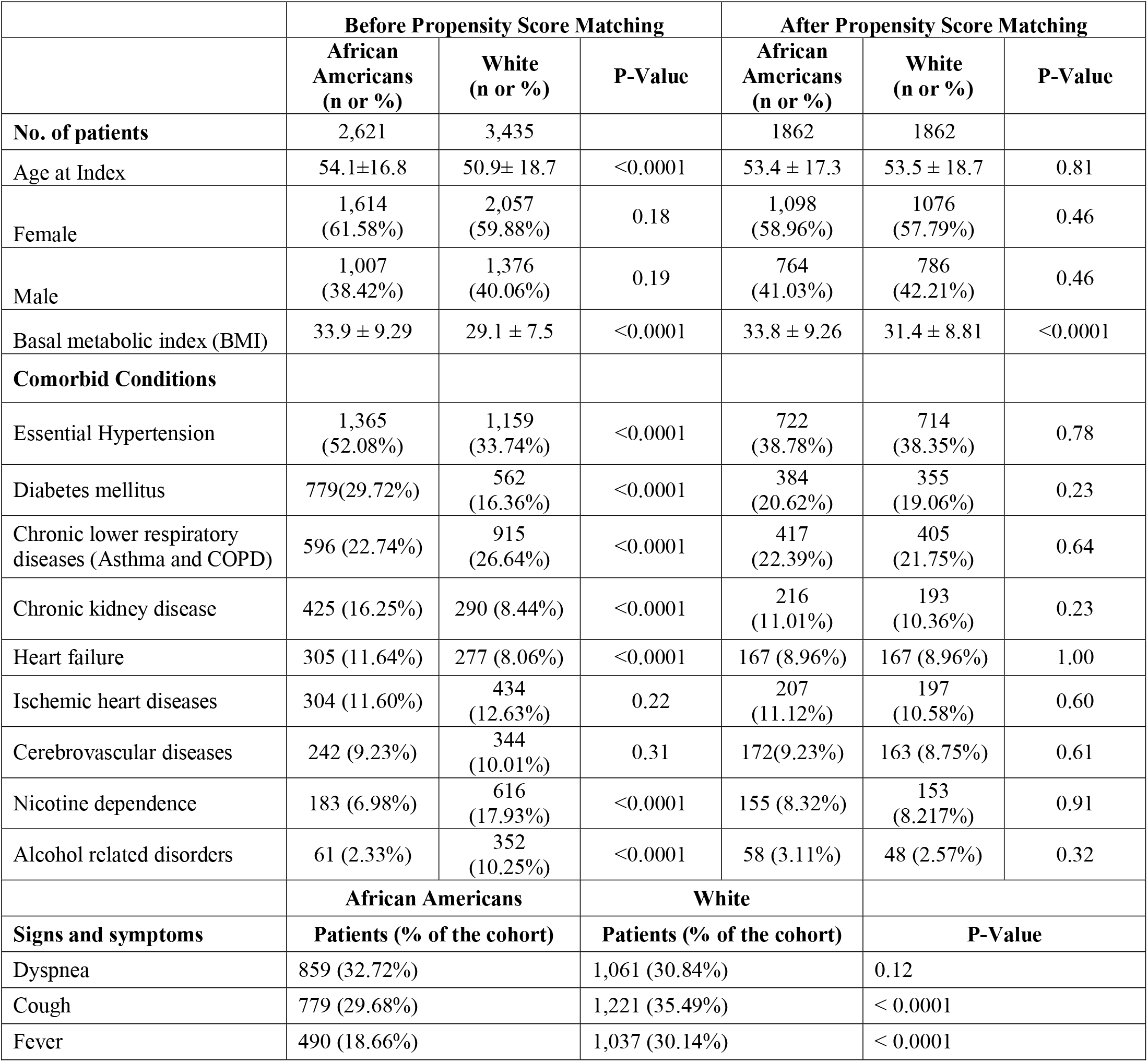
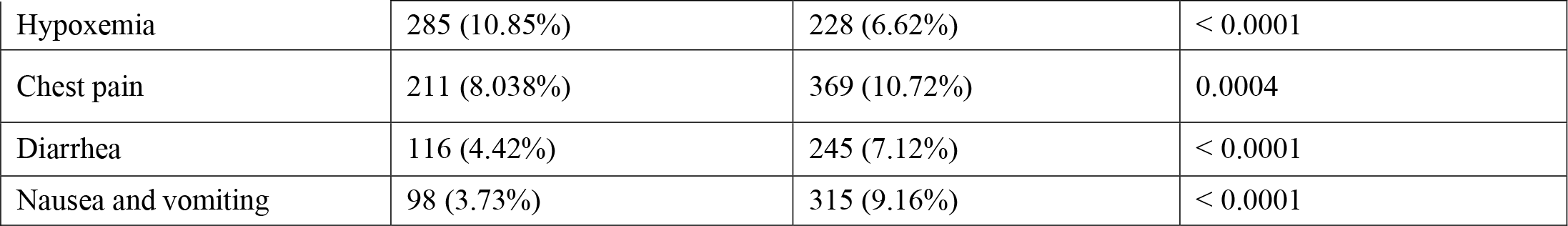
Comparison of patient characteristics and presenting signs and symptoms among COVID-19 African American and White patients. Demographics and comorbidities are compared before and after propensity matching of cohorts.

|  | Before Propensity Score Matching |  |  | After Propensity Score Matching |  |  |
| --- | --- | --- | --- | --- | --- | --- |
|  | African Americans<br>(n or %) | White<br>(n or %) | P-Value | African Americans<br>(n or %) | White<br>(n or %) | P-Value |
| **No. of patients** | 2,621 | 3,435 |  | 1862 | 1862 |  |
| Age at Index | 54.1±16.8 | 50.9± 18.7 | <0.0001 | 53.4 ± 17.3 | 53.5 ± 18.7 | 0.81 |
| Female | 1,614<br>(61.58%) | 2,057<br>(59.88%) | 0.18 | 1,098<br>(58.96%) | 1076<br>(57.79%) | 0.46 |
| Male | 1,007<br>(38.42%) | 1,376<br>(40.06%) | 0.19 | 764<br>(41.03%) | 786<br>(42.21%) | 0.46 |
| Basal metabolic index (BMI) | 33.9 ± 9.29 | 29.1 ± 7.5 | <0.0001 | 33.8 ± 9.26 | 31.4 ± 8.81 | <0.0001 |
| **Comorbid Conditions** |  |  |  |  |  |  |
| Essential Hypertension | 1,365<br>(52.08%) | 1,159<br>(33.74%) | <0.0001 | 722<br>(38.78%) | 714<br>(38.35%) | 0.78 |
| Diabetes mellitus | 779(29.72%) | 562<br>(16.36%) | <0.0001 | 384<br>(20.62%) | 355<br>(19.06%) | 0.23 |
| Chronic lower respiratory diseases (Asthma and COPD) | 596 (22.74%) | 915<br>(26.64%) | <0.0001 | 417<br>(22.39%) | 405<br>(21.75%) | 0.64 |
| Chronic kidney disease | 425 (16.25%) | 290 (8.44%) | <0.0001 | 216<br>(11.01%) | 193<br>(10.36%) | 0.23 |
| Heart failure | 305 (11.64%) | 277 (8.06%) | <0.0001 | 167 (8.96%) | 167 (8.96%) | 1.00 |
| Ischemic heart diseases | 304 (11.60%) | 434<br>(12.63%) | 0.22 | 207<br>(11.12%) | 197<br>(10.58%) | 0.60 |
| Cerebrovascular diseases | 242 (9.23%) | 344<br>(10.01%) | 0.31 | 172(9.23%) | 163 (8.75%) | 0.61 |
| Nicotine dependence | 183 (6.98%) | 616<br>(17.93%) | <0.0001 | 155 (8.32%) | 153<br>(8.217%) | 0.91 |
| Alcohol related disorders | 61 (2.33%) | 352<br>(10.25%) | <0.0001 | 58 (3.11%) | 48 (2.57%) | 0.32 |
|  | **African Americans** |  | **White** |  |  |  |
| **Signs and symptoms** | **Patients (% of the cohort)** |  | **Patients (% of the cohort)** | **P-Value** |  |  |
| Dyspnea | 859 (32.72%) |  | 1,061 (30.84%) | 0.12 |  |  |
| Cough | 779 (29.68%) |  | 1,221 (35.49%) | < 0.0001 |  |  |
| Fever | 490 (18.66%) |  | 1,037 (30.14%) | < 0.0001 |  |  |

|  |  |  |  |
| --- | --- | --- | --- |
| Hypoxemia | 285 (10.85%) | 228 (6.62%) | < 0.0001 |
| Chest pain | 211 (8.038%) | 369 (10.72%) | 0.0004 |
| Diarrhea | 116 (4.42%) | 245 (7.12%) | < 0.0001 |
| Nausea and vomiting | 98 (3.73%) | 315 (9.16%) | < 0.0001 |

**Figure 2:**
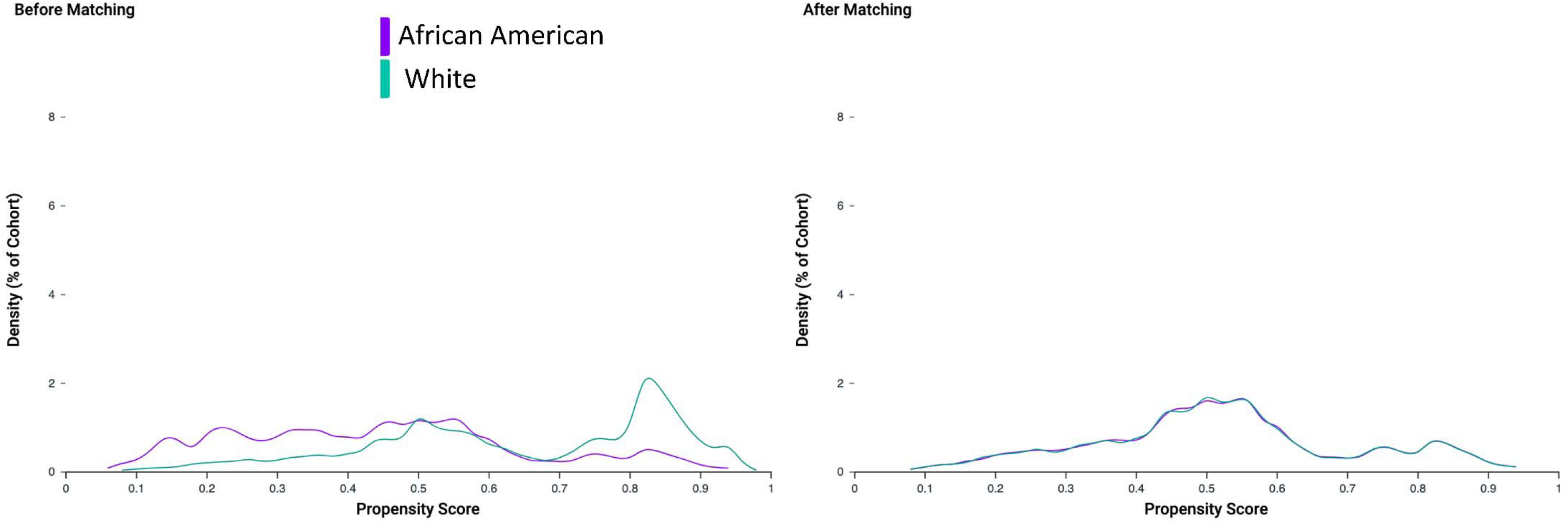
Propensity Score Density Graph before and after propensity score matching of the African-American and White cohort of patients.

The risk of 30-day mortality was significantly higher in both the unmatched (8.08% vs. 4.22%, RR 1.91, 95 % CI: 1.56,2.35, P <0.001) and matched black patients (6.92% vs. 5.20%, RR 1.33, 95 % CI: 1.03,1.72, P 0.02) compared to whites. Overall and age-wise subgroup analysis of mortality outcomes is described in Table 2 and Figure 3. A higher proportion of patients in the black cohort required hospitalization, invasive mechanical ventilation, and critical care services compared to the White cohort. Similarly, the prevalence of ARDS, AKI, requirement for CRRT, and septic shock were also higher in the black cohort (Table 2). Characteristics of non-survivors, patients requiring mechanical ventilation, and hospitalizations among black and White patients are compared in Supplementary Table 4.

**Table 2:** Outcomes among COVID-19 African American and White patients. Outcomes are compared before and after propensity score matching of cohorts.

|  | Before Propensity Score Matching |  |  |  |  | After Propensity Score Matching |  |  |  |  |
| --- | --- | --- | --- | --- | --- | --- | --- | --- | --- | --- |
|  | African Americans | White |  |  |  | African Americans | White |  |  |  |
| Outcomes | Patients with outcomes (Risk) | Patients with outcomes (Risk) | Risk Ratio (95% confidence interval) | Risk difference (95% confidence interval) | P-Value | Patients with outcomes (Risk) | Patients with outcomes (Risk) | Risk Ratio (95% confidence interval) | Risk difference (95% confidence interval) | P-Value |
| **Mortality** |  |  |  |  |  |  |  |  |  |  |
| **Overall** | 212/2,621 (8.08%) | 145/3,435 (4.22%) | 1.91 (1.56, 2.35) | 3.86% (2.62%,5.10%) | <0.001 | 129/1,862 (6.92%) | 97/1,862 (5.20%) | 1.33 (1.03,1.72) | 1.72% (0.19%,3.25%) | 0.02 |
| **Age ≤ 49** | 22/1,028 (2.14%) | 16/1,674 (0.96%) | 2.23 (1.18, 4.24) | 1.18% (0.18%,2.18%) | <0.001 | 12/754 (1.59%) | 10/754 (1.32%) | 1.2 (0.52, 2.76) | 0.26% (-0.94%,1.47%) | 0.67 |
| **Age 50-64** | 58/838 (6.92%) | 26/900 (2.89%) | 2.39 (1.52, 3.76) | 4.03% (1.99%,6.07%) | <0.001 | 33/550 (6.00%) | 15/550 (2.72%) | 2.2 (1.20,4.00) | 3.27 (0.86,5.67) | 0.01 |
| **Age ≥65** | 132/755 (17.48%) | 103/862 (11.94%) | 1.46 (1.15,1.85) | 5.53% (2.06%, 9.00%) | 0.0016 | 104/535 (19.43%) | 73/535 (13.64%) | 1.42 (1.08,1.87) | 5.79% (1.35%, 10.23%) | 0.01 |
| **Mechanical Ventilation** | 179/2,621 (6.82%) | 78/3,435 (2.27%) | 3.01(2.31,3.91) | 4.55% (3.47%,5.64%) | <0.001 | 118/1,862 (6.33%) | 50/1,862 (2.68%) | 2.36 (1.71,3.26) | 3.65% (2.32%,4.98%) | < 0.0001 |
| **Critical Care Services** | 419/2,621 (15.98%) | 179/3,435 (5.21%) | 3.06 (2.59,3.62) | 10.77% (9.18%,12.36%) | <0.001 | 263/1,862 (14.12%) | 134/1,862 (7.20%) | 1.96 (1.61,2.39) | 6.92% (4.96%,8.90%) | < 0.0001 |
| **Hospitalization** | 1,023/2,621 (39.03%) | 565/3,435 (16.45%) | 2.37 (2.17, 2.59) | 22.58% (20.34%,24.82%) | <0.001 | 684/1,862 (36.73%) | 402/1,862 (21.59%) | 1.70 (1.53,1.90) | 15.15% (12.37%,18.02%) | < 0.0001 |
| **Acute Respiratory Distress Syndrome** | 236/2,621 (9.01%) | 108/3,435 (3.14%) | 2.86 (2.29,3.57) | 5.86% (4.61%,7.10%) | <0.001 | 147/1,862 (7.89%) | 75/1,862 (4.03%) | 1.96 (1.49,2.56) | 3.87% (2.35%,5.38%) | < 0.0001 |
| **Acute Kidney Injury** | 361/2,621 (13.73%) | 167/3,435 (4.86%) | 2.83 (2.37,3.37) | 8.91% (7.41%,10.41%) | <0.001 | 234/1,862 (12.57%) | 107/1,862 (5.74%) | 2.18 (1.76,2.72) | 6.82% (4.98%,8.66%) | < 0.0001 |
| **Septic Shock** | 82/2,621<br>(3.12%) | 52/3,435<br>(1.51%) | 2.06<br>(1.46,2.91) | 1.61%<br>(0.83%,2.39%) | <0.001 | 46/1,862<br>(2.47%) | 29/1,862<br>(1.55%) | 1.58<br>(1.001,2.51) | 0.91%<br>(0.001%,1.81%) | 0.04 |
| **Continuous**<br>**Renal**<br>**Replacement**<br>**Therapy** | 50/2,621<br>(1.91%) | 21/3,435<br>(0.61%) | 3.12<br>(1.87,5.18) | 1.29%<br>(0.71%,1.88%) | <0.001 | 29/1,862<br>(1.56%) | 18/1,862<br>(0.96%) | 1.61<br>(0.89,2.89) | 0.59% (-<br>0.13%,1.30%) | 0.1 |

**Figure 3:**
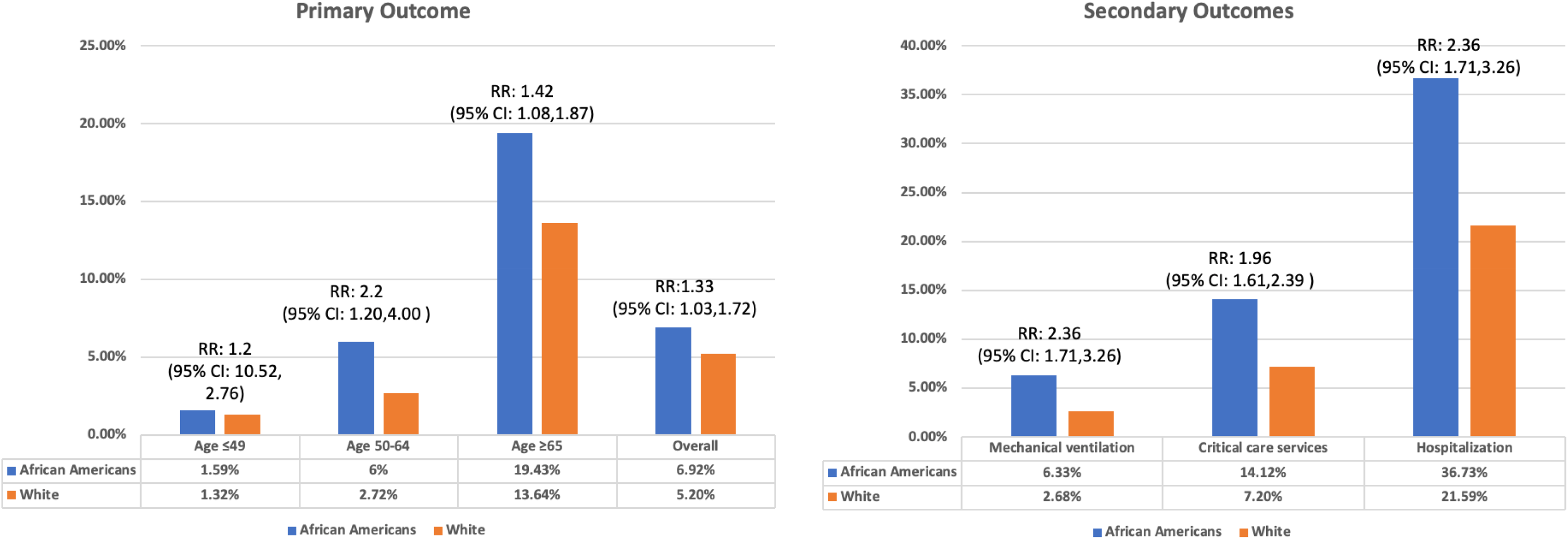
Comparison of Outcomes among African American and White patients after propensity score matching of cohorts. Mortality is compared in the overall cohort and in subgroups based on age. Secondary outcomes described in the overall cohort of patients.

## Discussion

Current data suggests worse COVID-19 outcomes in older patients with pre-existing HTN, DM, and cardiovascular diseases[3, 4], This is the first attempt to examine and understand the racial disparity in COVID-19 after disaggregating the effect of comorbidities. In the unmatched sample, we found a 91% increased risk of death in Blacks and a 137% higher need for hospitalization along with a greater than threefold higher need for mechanical ventilation. These findings go along with news media reports of drastically higher COVID-19 infection rates and mortality in Black patients in Chicago, Louisiana, and Michigan with up to 6-fold higher mortality in predominantly black counties compared to predominantly white ones[7-9].

A large part of these disparities in outcomes can be explained by the differences in high-risk characteristics between black and white patients (Unmatched RR for mortality 1.91 vs. matched RR 1.33). The cohorts derived from our data source are reasonably representative of the differences in these comorbidities vis-à-vis Black and White populations in the United States. It is well known that the incidence of HTN is exceptionally high amongst Black adults (>40%), and even if recognized, it is much less likely to be controlled[10-12]. Similarly, the prevalence of diabetes in our study cohort also mirrors known data about racial differences [13]. Obesity also disproportionately affects minorities and individuals from lower socioeconomic groups, particularly Blacks[14], Additionally, these comorbidities are not independent but interrelated, and having one disease also directly raises the hazard of developing others. A higher incidence of HTN and DM is associated with chronic kidney disease and, most likely, the higher prevalence of heart failure in the Black population[15, 16]. Moreover, the mortality from certain diseases is higher in Blacks than whites, despite being less or equally prevalent such as coronary heart disease and smoking[17, 18].

A sizeable incremental risk in Blacks persists even after accounting for an exhaustive list of comorbidities. Multiple factors likely interact in intricate ways to contribute to this disparity. It is possible that socioeconomic environments that promote poor health in the Black population likely have a critical influence on COVID-19 outcomes. Black communities still live in more segregated, closely spaced urban housing [19], and this may negatively impact the ability to practice social distancing. A disproportionate number of Blacks are employed in the service industry and in low paying jobs, which cannot be transitioned to working remotely during the COVID-19 pandemic[20]. These jobs also may lack protection from furloughs and lay-offs, requiring families to be forced to find work, which increases the chances of exposure. Blacks also face challenges in access to health care in the United States. Moreover, when they receive health care, it may not be equivalent to that for the white population[21], A greater number of Blacks lack health insurance and have a much higher rate of loss of insurance due to job/life circumstances[10, 22], Lack of access to health care can delay diagnosis and medical care in Blacks resulting in poor COVID-19 outcomes. Moreover, patient self-care, a behavior ranging from compliance with preventive measures to seeking health care, is perceived as lacking in Blacks [21].

Other potential etiologies also need to be explored – the angiotensin-converting enzyme 2 (ACE-2) receptor is responsible for COVID 19 virus entry into host cells and also has been hypothesized to play a part in the lung injury. The frequency of ACE-2 variants and their expression may explain susceptibility and severity of disease in different races, and a wide variety of allele frequency differences have already been shown in Asians, Eurasians and African populations[23]. There are also genetic polymorphisms for clotting factors, especially Fibrinogen in the Black population, which may increase thrombosis[24]. There are signals that thromboembolic complications are seen with sicker COVID-19 patients, and a link between this and increased mortality in Black patients may be a potential area for investigation.

Our analysis carries the limitations of using an EMR database despite the ability of TriNetX to aggregate the data directly from the EMRs in a real-time fashion minimizing the risk of data collection errors. Patient counts were rounded up to the nearest 10 in our analysis to guard Protected Health Information (PHI). Rounding may influence measures of association results for small cohorts and infrequent outcomes; however, most of our outcomes have a relatively large number of patients. Exposure history, incubation time, and dynamic changes in patients’ clinical condition could not be estimated. Socioeconomic factors, geographical variations, delivery, and access to health care during COVID-19 were beyond the scope of our study; however, they have a significant influence on differences in high-risk characteristics and outcomes for COVID-19. We did not include other minorities in our study because of the relatively small sample size.

In conclusion, our study provides evidence of poor outcomes among Black patients affected by COVID-19. Our results indicate the role of disproportionate comorbidities in the Black population but also shows an unexplained significant residual difference that persists after careful propensity matching. This highlights the role socioeconomic factors and healthcare disparities have played in the COVID-19 pandemic and the need for a vast improvement in the care of Black population.

## Data Availability

Supplementary Material

## Acknowledgment

We acknowledge the West Virginia Clinical and Translational Science Institute to provide us access, and training to the TriNETX global healthcare network. We also acknowledge the TriNETX (Cambridge, MA, USA) healthcare network for design assistance to complete this project.The authors received no financial support or grants for the research, authorship, and publication of this article.

Ahmad Khan, Arka Chatterjee, and Shailendra Singh declare that they have no conflict of interest.

## Data availability

Data is available to member Health Care Organizations on the TriNetX https://www.trinetx.com/ research network platform. Data aggregated directly from the electronic health records systems of the participating HCOs is provided a real-time fashion on the TriNetX cloud-based platform. Datasets for individual patient level data can be requested from TriNetX by participating Health Care Organizations. Codes for creating cohort of COVID-19 patients races are included in methods. Race is coded to HL7 version 3 administrative standards. Codes for other characteristics, laboratory values, and outcomes are included in the supplementary.

## Supplementary

Supplementary Methods: Data Source, data quality checks, mapping, and coding system to present data.

Supplementary Table 1: Search query using ICD-10 and CPT codes used to define outcomes of the study.

Supplementary Table 2: TriNetX (TNX) laboratory or Logical Observation Identifiers Names and Codes (LOINC) to capture laboratory values.

Supplementary Table 3: Comparison of Laboratory findings in African American and White patients after diagnosis of COVID-19.

Supplementary Table 4: Comparison of characteristics in patients with mortality, mechanical ventilation, and hospitalizations among African Americans and Whites.

## References

1. COVID-19 Dashboard by the Center for Systems Science and Engineering (CSSE) at Johns Hopkins University (JHU). [cited 2020 April 19th]; Available from: https://coronavirus.jhu.edu/map.html.

2. Garg S, K.L., Whitaker M, et al, Hospitalization Rates and Characteristics of Patients Hospitalized with Laboratory-Confirmed Coronavirus Disease 2019—COVID-NET, 14 States March 1–30, 2020. MMWR Morb Mortal Wkly Rep 2020;69:458–464.

3. Guo, T., et al., Cardiovascular Implications of Fatal Outcomes of Patients With Coronavirus Disease 2019 (COVID-19). JAMA Cardiol, 2020.

4. Shi, S., et al., Association of Cardiac Injury With Mortality in Hospitalized Patients With COVID-19 in Wuhan, China. JAMA Cardiol, 2020.

5. Carnethon, M.R., et al., Cardiovascular Health in African Americans: A Scientific Statement From the American Heart Association. Circulation, 2017. 136(21): p. e393-e423.

6. Race & Ethnicity. United States Census Bureau, [cited 2020 April 26]; Available from: https://www.census.gov/mso/www/training/pdf/race-ethnicity-onepager.pdf.

7. Thebault R, B.T.A., Williams V The coronavirus is infecting and killing black Americans at an alarmingly high rate, in Washington Post. 2020.

8. Reyes C, H.N., Gutowski C, St Clair S, Pratt G, Chicago’s coronavirus disparity: black Chicagoans are dying at nearly six times the rate of white residents, data show, in Chicago Tribune. 2020.

9. Deslatte, M., Louisiana data: virus hits blacks, people with hypertension, in AP News. 2020.

10. in Health, United States, 2015: With Special Feature on Racial and Ethnic Health Disparities. 2016: Hyattsville (MD).

11. Mills, K.T., et al., Global Disparities of Hypertension Prevalence and Control: A Systematic Analysis of Population-Based Studies From 90 Countries. Circulation, 2016. 134(6): p. 441–50.

12. Howard, G., et al., Racial and geographic differences in awareness, treatment, and control of hypertension: the REasons for Geographic And Racial Differences in Stroke study. Stroke, 2006. 37(5): p. 1171-8.

13. Menke, A., et al., prevalence of and Trends in Diabetes Among Adults in the United States, 1988-2012. JAMA, 2015. 314(10): p. 1021-9.

14. Abraham, P.A., et al., Obesity and african americans: physiologic and behavioral pathways. ISRN Obes, 2013. 2013: p. 314295.

15. Lopes, A.A., End-stage renal disease due to diabetes in racial/ethnic minorities and disadvantaged populations. Ethn Dis, 2009.19(1 Suppl 1): p. S1–47-51.

16. Bahrami, H., et al., Differences in the incidence of congestive heart failure by ethnicity: the multi-ethnic study of atherosclerosis. Arch Intern Med, 2008. 168(19): p. 2138-45.

17. Safford, M.M., et al., Association of race and sex with risk of incident acute coronary heart disease events. JAMA, 2012. 308(17): p. 1768-74.

18. African Americans and Tobacco Use. November 18, 2019 [cited 2020 April 26th]; Available from: https://www.cdc.gov/tobacco/disparities/african-americans/index.htm.

19. Sohn, H., Racial and Ethnic Disparities in Health Insurance Coverage: Dynamics of Gaining and Losing Coverage over the Life-Course. Popul Res Policy Rev, 2017.36(2): p. 181–201.

20. Toossi, E.R.a.M. Blacks in the labor force. February 2018 [cited 2020 April 24]; Available from: https://www.bls.gov/careeroutlook/2018/article/blacks-in-the-labor-force.htm?viewfull.

21. National Research Council (US) Panel on Race, Ethnicity, and Health in Later Life, in Understanding Racial and Ethnic Differences in Health in Late Life, A.N. Bulatao RA, Editor. 2004, National Academies Press (US): Washington (DC).

22. Edward R. Berchick, J.C.B., and Rachel D. Upton. Health Insurance Coverage in the United States: 2018. 2019 [cited 2020 April 24]; Available from: https://www.census.gov/content/dam/Census/library/publications/2019/demo/p60-267.pdf.

23. Cao, Y., et al., Comparative genetic analysis of the novel coronavirus (2019-nCoV/SARS-CoV-2) receptor ACE2 in different populations. Cell Discov, 2020. 6: p. 11.

24. Kotze, RC, et al., Genetic polymorphisms influencing total and gamma’ fibrinogen levels and fibrin clot properties in Africans. Br J Haematol, 2015. 168(1): p. 102–12.

